# MultiECGNet: A novel deep learning-based multi-format ensemble method for image-based electrocardiographic diagnosis of atrial fibrillation

**DOI:** 10.1101/2025.06.21.25330061

**Authors:** Justin Phan, George Parker, Nigel Lovell, Rajesh N Subbiah, Jamie I Vandenberg, Adam P. Hill, Ahmadreza Argha

## Abstract

**Aim:** To evaluate the performance of an ensemble classifier, MultiECGNet, using multi-format electrocardiographic (ECG) images for the diagnosis of atrial fibrillation (AF), and to compare its performance with a signal-based deep learning model.

**Methods:** An ensemble of ECG classifiers was developed using four models derived by truncating the pre-trained EfficientNet B3 model at different feature extraction layers. Transfer learning was employed to train the ensemble on the publicly available PTB-XL dataset for AF detection. External validation was performed using the Chinese Physiological Society Signal Challenge 2018 dataset. ECG samples were converted into two image formats (2x6 and 4x3), and performance was evaluated across same-format, cross-format, and mixed-format classification tasks.

**Results:** The ensemble classifier detected AF in the external validation dataset with an accuracy of 0.95 and F1 score of 0.87, which was comparable to a signal-based model (F1-score: 0.87 vs 0.83) and outperformed a single EfficientNet-B3 model (F1-score: 0.87 vs 0.71). Training on one ECG format and testing on a different format resulted in reduced performance (F1 score: 0.66-0.71). However, training on a dataset containing a balanced mix of both formats improved performance compared to same-format training alone (F1-score: 0.86 vs 0.87).

**Conclusion:** The proposed image-based ensemble classifier demonstrated comparable performance to a strong signal-based model for AF detection. While cross-format generalization posed a challenge, incorporating multiple ECG formats during training mitigated this limitation and improved model robustness.

## Introduction

Electrocardiography (ECG) is a widely used, non-invasive diagnostic tool that records the electrical activity of the heart. ECG analysis is a critical diagnostic method for detecting a range of cardiovascular conditions, such as arrhythmias, myocardial infarction, and other heart abnormalities[1]. One important use of ECG analysis is in the evaluation and management of transient ischemic attacks (TIA). Atrial fibrillation (AF)–related ischemic strokes and TIAs are leading contributors to recurrent stroke risk, which can be significantly reduced through appropriate oral anticoagulant therapy. Traditionally, ECG analysis has relied on expert interpretation and rule-based algorithms. However, the emergence of deep learning has introduced a paradigm shift in the analysis of ECG data. While most deep learning applications in ECG analysis have utilised one-dimensional convolution neural networks (1-D CNNs) applied to raw digital signals, such data are not always readily available, particularly in retrospective studies. Therefore, the use of two-dimensional CNNs (2-D CNNs) to analyse ECG images is gaining traction. For example, Sangha *et al*. recently showed that a CNN trained on ECG images could predict echocardiographic left ventricular dysfunction with a sensitivity of 89% and specificity of 77%[2] . This image-based approach is particularly promising for application to large repositories of historical ECGs. The ability to leverage historical ECG archives is particularly advantageous in the study of rare conditions, where prospective data collection would otherwise require extensive time and resources.

Prior work has shown that image-based 2D-CNNs can perform comparably to signal-based 1D-CNNs, particularly when applied to smaller datasets. This is partly due to the use of transfer learning, where 2D-CNNs pre-trained on large image datasets (such as ImageNet[3] ) are fine-tuned on smaller ECG image datasets. This approach enables the model to leverage previously learned visual features, improving performance and reducing the risk of overfitting when labelled ECG data is limited. For example, Wu *et al.* used AlexNet, a 2D-CNN architecture, to classify normal versus abnormal QRS complexes based on ECG images, achieving an accuracy of 98% in detecting abnormalities[4] . However, for more complex classification tasks, such as diagnosing arrhythmias like AF, there remains limited comparative research between image-based and signal-based deep learning approaches.

Using ECG images for machine learning, however, also presents challenges compared to digital signal-based approaches. Notably, neural networks may inadvertently learn from non-physiological features, such as the layout of ECGs, which must be considered in model design and evaluation. To our knowledge, only one study has evaluated the performance of an image-based deep learning model trained to classify ECG images with multiple diagnostic labels (e.g., left bundle branch block, right bundle branch block and AF) across different ECG formats (2x6 and 4x3 layouts)[2]. On a small external validation set of 64 ECG images, the model showed good performance, with F1 scores ranging from 0.73 to 0.98.

A major concern when using deep learning on small datasets is the risk of overfitting. Ensemble learning, a technique that combines predictions from multiple models, can mitigate this risk and has been shown to improve performance compared to using a single network[5]. Accordingly, in this study, we aimed to test the hypothesis that ensemble learning can enhance the performance of a moderately sized ECG-image based deep learning model (trained on thousands of ECGs) for the identification of atrial fibrillation, regardless of ECG format used to present the data. We developed a novel ensemble of truncated networks based on the publicly available EfficientNet B3 architecture [6] and compared its performance to that of a single EfficientNet B3 model, as well as to a signal-based deep learning approach for classifying ECGs with and without AF. Our ensemble model, called *MultiECGNet*, demonstrated superior performance compared to a single EfficientNet B3 and comparable performance to a high-performing signal-based deep learning model, while maintaining robust classification across the two most common ECG image formats.

## Methods

### Data acquisition and pre-processing

We used data from the publicly available PTB-XL dataset[7], accessible via PhysioNet (physionet.org/content/ptb-xl/1.0.3/). The dataset comprises 21,799 12-lead ECGs collected from 18,869 patients that were annotated by up to two cardiologists. Among these, 5,071 ECGs were annotated as showing AF.

The raw ECG signal data was converted into electrocardiographic images using the Python ecg-plot function[8] . By default, this function renders the ECG in a 6x2 lead format, displaying 5 seconds of data per lead. We modified the function to also generate images in a 3x4 lead layout, where each lead displays 2.5 seconds of data. Examples of both ECG formats are presented in Figure 1. All images were resized to 300x300 pixels for compatibility with the EfficientNet B3 CNN.

**Figure 1.**
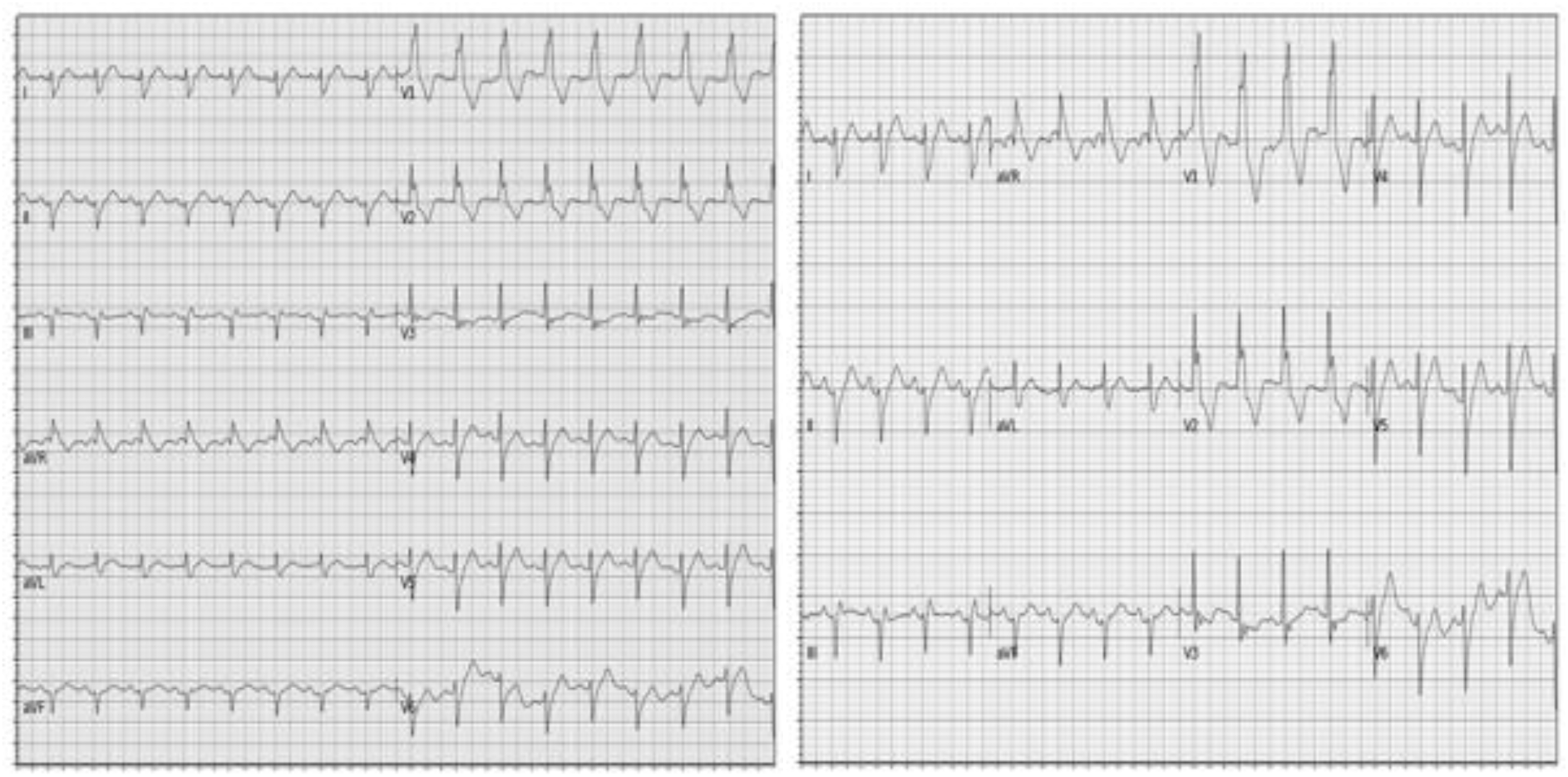
Sample 12-lead ECG images generated using ecg-plot function, shown in 2x6 format (left) and 3x4 format (right) and reduced to 300 x 300 pixels to match the input required for the EfficientNetB3 algorithm

The labels atrial fibrillation (AF) and not atrial fibrillation (not AF) were used for binary classification. The PTB-XL dataset was separated into three subsets using a stratified split to preserve class distribution; 60% for training, 20% for validation and 20% as a holdout set for model testing. External validation was subsequently performed using the publicly available Chinese Physiological Signal Challenge (CPSC) dataset, which contains 6,877 12-lead ECG recordings of which 1,094 were annotated as atrial fibrillation[9] .

### Model Architecture

#### EfficientNet

EfficientNet is a family of 2-D CNNs, developed by Google Research, designed to achieve high accuracy while optimising computational efficiency [6] . EfficientNet B3, the fourth model in this family, offers a strong balance between performance and computing resource consumption. It requires input images of 300x300 pixels, includes 388 layers, and contains over 10 million trainable parameters. Initially trained on the ImageNet dataset, which includes over 1,000 object categories, EfficientNet B3 can be fine-tuned for downstream tasks using transfer learning [10] making it well-suited for applications such as ECG image classification.

### Model Variants

Four ensemble-based classification strategies were developed and compared:

#### 1-Snapshot Ensemble

The first approach used a snapshot ensemble (Figure 2A), in which a single model is saved at different stages of training. A single EfficientNet B3 model was fine-tuned for 500 epochs using cosine annealing, as described by Huang *et al.* [11] , with the learning rate undergoing 50 annealing cycles. At the end of each cycle, a snapshot of the model was saved, resulting in 50 distinct models.

**Figure 2.**
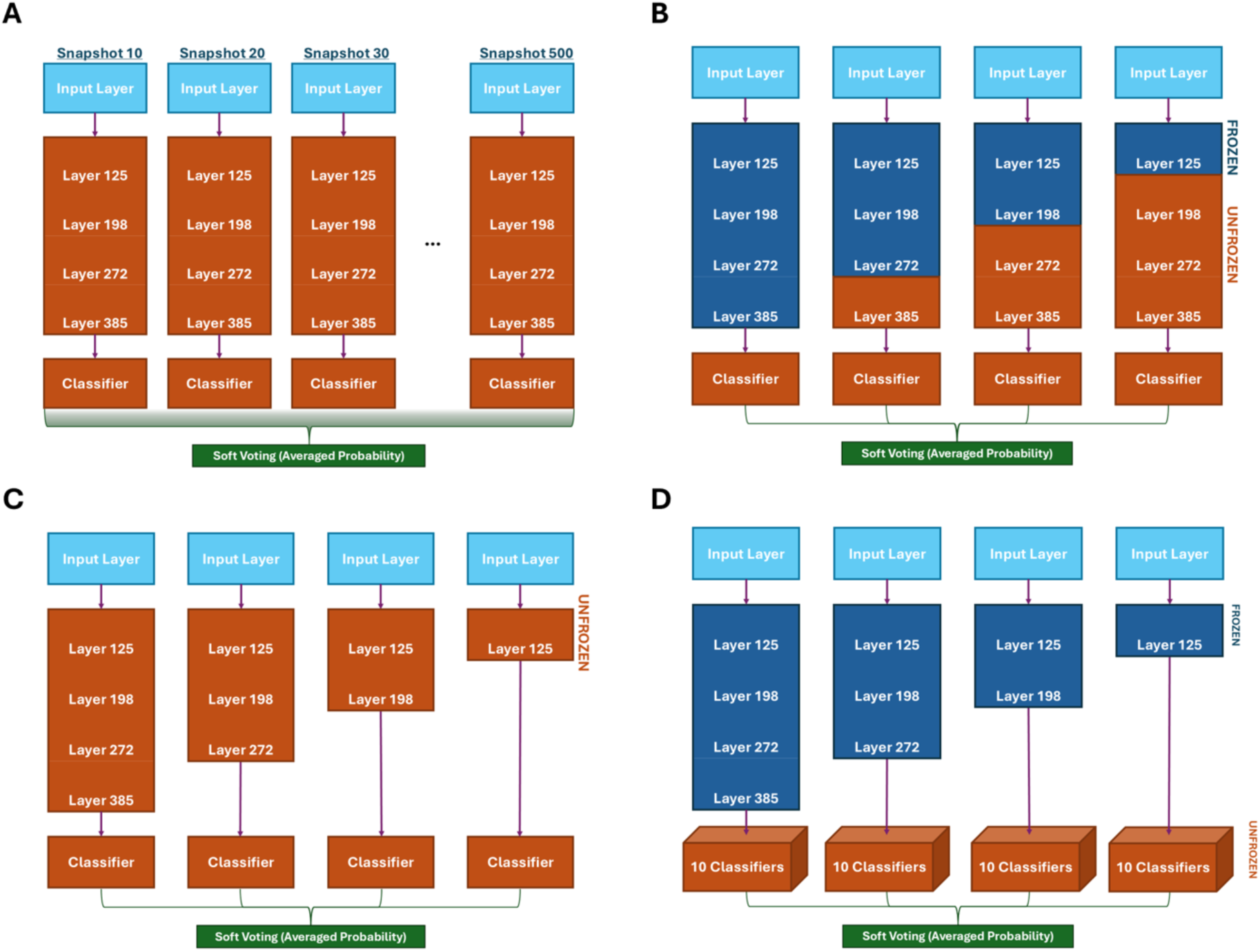
The four different training strategies employed to create the model ensembles; (A) the Snapshot Ensemble, (B) the FreezNet strategy using pretrained ImageNet weights, (B) the RetrainNet strategy and (D) the DiverseNet strategy using the retrained weights from RetrainNet. Blue signifies the frozen layers of the network, while orange signifies unfrozen layers that are retrained

#### 2-Partially Frozen Fine-Tuning (FreezNETs)

This fine-tuning method was adapted from the soft cut approach described by Hansun *et al.* [12] . In this approach, EfficientNet B3 models were initialised with pretrained weights. All layers prior to a selected point in the network were frozen (i.e., weights not updated during training), while the subsequent layers remained trainable (Figure 2B). This enabled fine-tuning of deeper features without altering the lower-level representations learned during pretraining. The pretrained classification layers were removed and new classifiers were attached to accommodate the classification task. These classifiers consisted of an average pooling layer, two fully connected layers (with randomly selected sizes from {32, 64, 128, 256, 512, 1024}), a dropout layer, and a final classification layer. The selected points for freezing were at layers 125, 198, 272 or 385—corresponding to the end of key squeeze-and-excitation blocks, which play a central role in the network’s feature extraction hierarchy.

#### 3-Full Fine-Tuning (RetrainNETs)

In this setup, 4 truncated variations of the pre-trained EfficientNet B3 were created by extracting features from the same four layers chosen in the FreezNets approach; 125, 198, 272 or 385, but here all layers after the truncation were removed (Figure 2C). Remaining layers were unfrozen (i.e., weights allowed to update during training) and a new classifier with similar randomized architecture to the classifiers described in the FreezNets was attached to each of the models. The purpose of this approach is to diversify the models in the ensemble by altering model depth and extracting different features for each classifier.

#### 4-Feature-Based Ensemble with Diverse Classifiers (DiverseNETs)

Based on the fully fine-tuned RetrainNETs, classification heads were removed, and the remaining networks were frozen to serve as fixed feature extractors (Figure 2D). Hansun *et al.* proposed using an ensemble classifier consisting of ten distinct networks to generate a probability distribution for each class output [12] . Adapting this technique for each of our frozen feature extractors, ten separate classifiers were trained with similar architectures and randomised sizes to the classifiers described in the FreezNet approach. The resulting output of the ensemble consisted of 40 probability scores for the classification of atrial fibrillation. This setup introduced diversity into the ensemble, enhancing robustness and generalisability.

### Soft Voting for Final Classification

A soft voting system was used to generate the final classification output from the ensemble models, where the predicted class was determined by averaging the probability scores from each model and selecting the class with the highest average probability.

### DigitalNet

For benchmarking purposes, our imaged-based classifier was compared to a model trained on raw ECG signal data. We used the neural network developed by Cai *et al.* [13]. Their model was originally trained on 16,557 samples from the CPSC and local 12-lead ECG datasets, achieving high performance over 99% accuracy, sensitivity, specificity and F1-score for binary classification between AF and normal rhythm. In our study, this model was retrained from scratch using the PTB-XL dataset, following the original training pipeline provided by the authors.

### Training Parameters and Hardware

We employed transfer learning by initialising model weights with pretrained EfficientNet B3 weights. The models were trained using the Adam optimiser [14] with a learning rate of1x10^-5^ and a mini-batch size of 32. Early stopping was applied, terminating raining if the validation loss did not improve over 10 consecutive epochs. Training was performed using an NVIDIA RTX A6000 GPU with 48 GB of RAM. All code was developed in Python 3.11.9 using Keras 2.9.0 as the frontend with a TensorFlow 2.9.1 backend.

### Assessing the impact of ECG lead formats on image-based classification

Firstly, to assess the effect of cross-format classification, all image-based models were trained using either 2x6 or 4x3 image formats from the PTB-XL dataset, with external validation performed using the opposite format from the CPSC dataset. Secondly, to evaluate the effect of combining lead formats, a training set was created by including both 2x6 and 3x4 images for each ECG from the PTB-XL dataset, effectively duplicating each ECG in two formats. External validation was then performed on a similarly combined CPSC dataset. Finally, to investigate the effect of training on a mixture of lead formats without duplication, a training set was generated by including an equal proportion of 2x6 and 3x4 images from the PTB-XL dataset, with each ECG represented only once. External validation was again performed on a similarly mixed-format CPSC dataset.

A class activation mapping technique, called gradient-weighted class activation map (GradCAM) was used to visualise the regions of the ECG images that most influenced the classification decisions [2], [15]. The GradCAM heatmaps were created using a weighted average of the probability scores across the models in the 40-classifier RetrainNets ensemble.

### Excluded Samples due to signal artefacts

A total of 17 samples were excluded from the PTB-XL dataset as they caused errors when processed by the DigitalNet. Each of these samples was manually reviewed by a cardiac electrophysiologist (Author J.P.), who confirmed that in every case, one or more leads demonstrated artefact consistent with poor electrode contact.

### Performance metrics

Each model’s performance was evaluated using the metrics of accuracy, precision, recall, and F1-score.

## Results

The performance of the various models on the classification task of atrial fibrillation vs not atrial fibrillation using a single ECG lead format (2x6) is shown in Table 1. The single EfficientNet B3 model demonstrated the lowest performance among all models tested, with particularly poor results in precision and F1-score. The DigitalNet had an intermediate performance with an F1-score of 0.83. The Snapshot Ensemble, RetrainNETs and DiverseNETs delivered the highest performance, each attaining an F1-score of 0.87. Accuracy, precision and recall were comparable across these three models.

**Table 1.** Classification of atrial fibrillation versus not atrial fibrillation for the 2x6 ECG lead format. Models were trained on the PTB-XL dataset, and the classification results are shown with external validation on the CPSC dataset. TP = True Positive, FP = False Positive, TN = True Negative, FN = False Negative.

| Method | EfficientNetB3 | SnapshotEnsemble | FreezNETs | RetrainNETs | DiverseNETs | DigitalNet |
| --- | --- | --- | --- | --- | --- | --- |
| Atrial fibrillation | 1094 | 1094 | 1094 | 1094 | 1094 | 1094 |
| Not atrial fibrillation | 5772 | 5772 | 5772 | 5772 | 5772 | 5772 |
| TP | 770 | 1001 | 917 | 1016 | 1014 | 997 |
| FP | 312 | 213 | 374 | 238 | 236 | 310 |
| TN | 5460 | 5559 | 5398 | 5534 | 5536 | 5462 |
| FN | 324 | 93 | 177 | 78 | 80 | 97 |
| Accuracy | 0.91 | 0.95 | 0.92 | 0.95 | 0.95 | 0.94 |
| Precision | 0.711 | 0.82 | 0.71 | 0.81 | 0.81 | 0.76 |
| Recall | 0.70 | 0.92 | 0.84 | 0.93 | 0.93 | 0.91 |
| F1-score | 0.71 | 0.87 | 0.77 | 0.87 | 0.87 | 0.83 |
| ROC AUC | 0.94 | 0.98 | 0.95 | 0.98 | 0.98 | 0.96 |

### Impact of ECG format

The results of training and validation using various ECG formats, as described above, are showed in Tables 2 and 3. The results presented correspond to the retrainNETs. Table 2 illustrates the decline in performance with cross-format classification with F1 scores of 0.66-0.71 compared to an F1 score of 0.87 with same-format classification. A marked reduction was also seen with precision and recall scores. Table 3 illustrates that there is a significant improvement in classification performance when the training dataset contains both ECG formats, in either a combined dataset (each ECG represented twice in 4x3 and 2x6 formats) or a mixed dataset (each ECG represented once with a 50-50 split of ECGs in 4x3 and 2x6 formats), with F1 scores of 0.86. All other performance metrics were similar between both training approaches.

**Table 2.** Cross-format classification results when models were trained on either 4x3 or 2x6 image formats.

| **Method** | **4x3 training with 2x6 validation** | **2x6 training with 4x3 validation** |
| --- | --- | --- |
| Atrial fibrillation | 1094 | 1094 |
| Not atrial fibrillation | 5772 | 5772 |
| TP | 883 | 581 |
| FP | 499 | 96 |
| TN | 5273 | 5676 |
| FN | 211 | 513 |
| Accuracy | 0.9 | 0.91 |
| Precision | 0.64 | 0.86 |
| Recall | 0.81 | 0.53 |
| F1-score | 0.71 | 0.66 |
| ROC AUC | 0.93 | 0.96 |

**Table 3.** Classification performance when training was performed on a combined dataset (comprising ECGs in both 4x3 and 2x6 formats with each ECG represented twice) and mixed datasets (comprising a random mixture of ECGs in 4x3 and 2x6 formats with each ECG represented once).

|  | **Combined training** | **Mixed Training** |
| --- | --- | --- |
| Atrial fibrillation | 2188 | 1094 |
| Not atrial fibrillation | 11544 | 5772 |
| TP | 1977 | 974 |
| FP | 437 | 185 |
| TN | 11107 | 5587 |
| FN | 211 | 120 |
| Accuracy | 0.95 | 0.96 |
| Precision | 0.82 | 0.84 |
| Recall | 0.90 | 0.89 |
| F1-score | 0.86 | 0.86 |
| ROC AUC | 0.97 | 0.97 |

### Training with combined and mixed datasets

To further explore the performance decline with cross-format training, the probability values for atrial fibrillation classification obtained from RetrainNETs using different ECG image formats (figure 3). This figure shows how individual models within the ensemble contributed to the final classification result. Cross-format classification, as shown in panels (C) and (D), demonstrated the poorest performance with the largest number of incorrect classifications and overall lower probability values compared to either same-format classification (panel A) or classification with mixed training (panel B), which had similar probability values.

**Figure 3.**
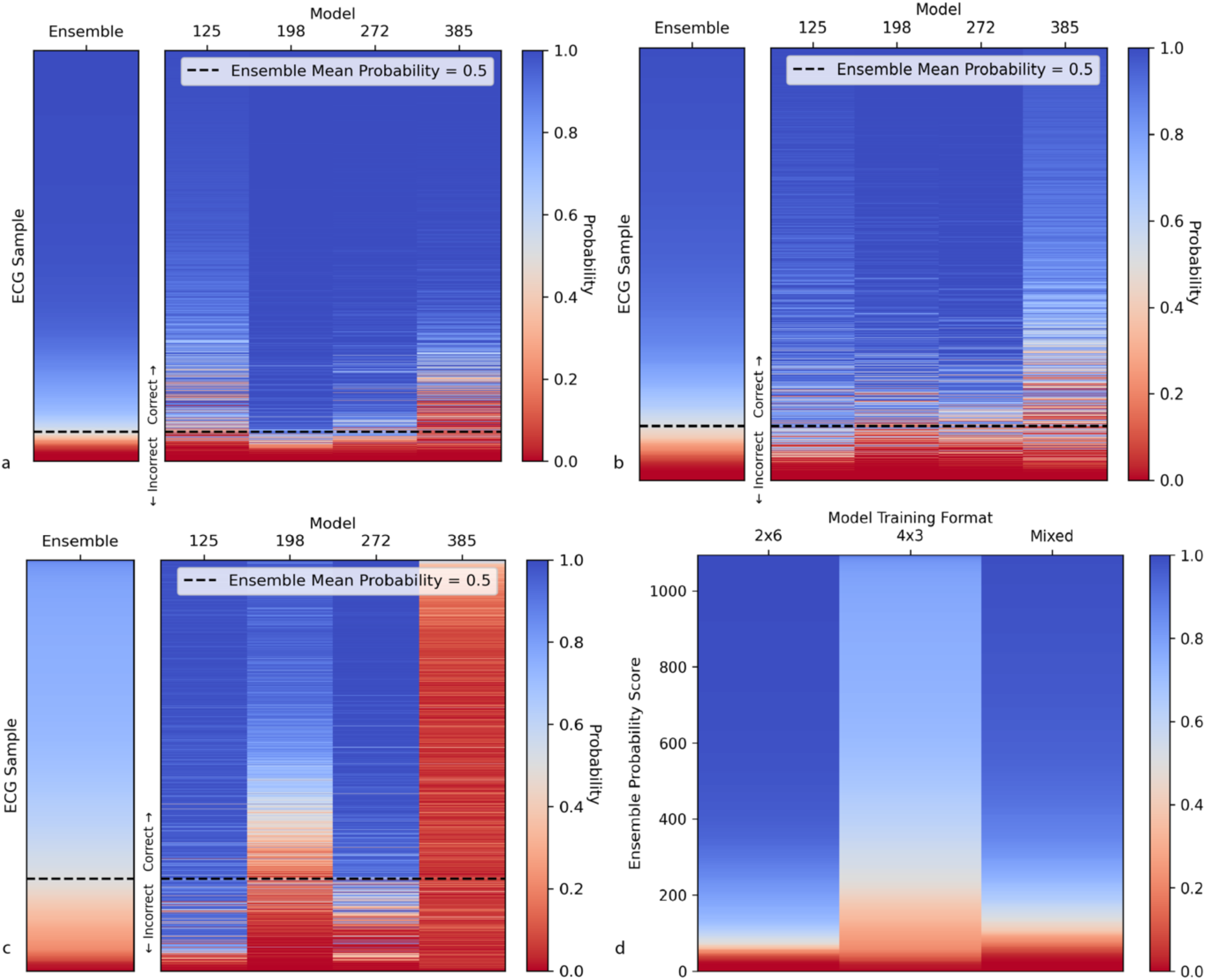
Panels (A), (B), and (C) show the probability values of the individual models from the RetrainNETs, along with the overall ensemble probability. Black dotted lines indicate the ensemble decision threshold of 0.5; probabilities above this line correspond to an ensemble classification of AF. Panel (A) shows results from training and testing on ECGs in the 2x6 format (same-format classification). Panel (B) shows results from training on a mixed ECG dataset and testing on the 2x6 format. Panel (C) shows the results of training on a 4x3 ECG format with cross-format (2x6) classification. Panel (D) presents the ensemble probability scores across all validation samples using each of the three training strategies: 2×6, 4×3, and mixed formats, with validation performed on the 2×6 format.

Notably, the full EfficientNet model within the ensemble (left most element in Figure 3C) demonstrated low confidence and made incorrect classifications during cross-format classification (panel (C)). We therefore tested the ensemble performance after removing the 385 layer component (Table 4). This removal led to a decline in overall performance compared to the four-model ensemble, with the F1 score decreasing from 0.71 to 0.64 (see Table 4).

**Table 4.** Cross-format classification performance after removing the poorly performing model 385.

| Model | 125 | 198 | 272 | Ensemble |
| --- | --- | --- | --- | --- |
| TP | 1015 | 697 | 925 | 916 |
| FP | 79 | 333 | 1250 | 815 |
| TN | 4043 | 5439 | 4522 | 4957 |
| FN | 1729 | 397 | 169 | 178 |
| Accuracy | 0.74 | 0.89 | 0.79 | 0.86 |
| Precision | 0.37 | 0.68 | 0.43 | 0.53 |
| Recall | 0.93 | 0.64 | 0.85 | 0.84 |
| F1-score | 0.53 | 0.66 | 0.57 | 0.65 |

### GradCAM

Figure 4 shows the GradCAM outputs for a single example ECG showing atrial fibrillation. The GradCAM images were generated by weighting the intensities from the individual models within the ensemble according to each model’s predicted probability, and then overlaying these weighted activations onto the ECG image. Panel (A) shows the activation map for a 2x6 ECG image with RetrainNETs trained on a dataset of 2x6 images. Panel (B) displays the activation map from the ensemble trained on 4x3 images. Panel (C) shows the activation map from the ensemble trained on the mixed dataset, where each ECG was represented once. Subsequently, the ECG images were divided into 10x10 pixel segments. Using grayscale thresholding, the ECG image was segmented into ‘signal’ and ‘no signal’ regions (middle panel). Finally, a histogram was generated to compare the frequency of activations within the ‘signal area’ compared to ‘non-signal’ area. This figure shows that the largest number of activations in the ‘no signal’ region occurred with cross-format classification (panel B), which is congruent with the lower confidence values seen in figure 3.

**Figure 4.**
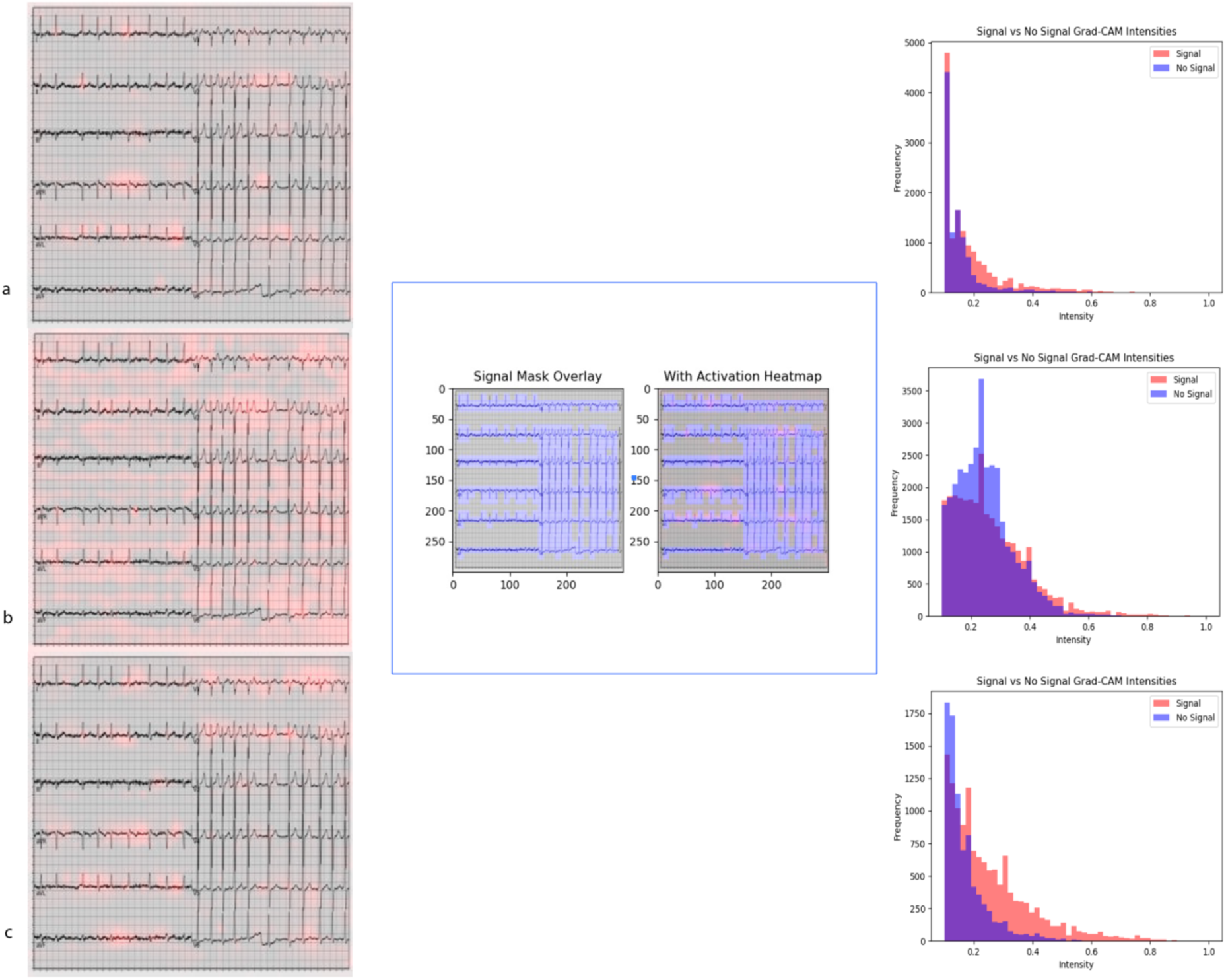
Weighted GradCAM maps of a single ECG example demonstrating atrial fibrillation. Panel (A) shows the activation map from same-format training, panel (B) presents the results from cross-format training, and panel (C) displays the results from mixed-format training.

## Discussion

In this study, we demonstrate that a novel ensembling approach can enhance the performance of image-based ECG analysis to a level comparable to that of a 1-D CNN trained on digital signals. For AF classification, our ensembles of neural networks, including RetrainNETs, Snapshot Ensemble, and DiverseNETs, achieved performance that was comparable to a high-performing digital signal-based model, and clearly outperformed a single EfficientNet B3 model. Previous work by Alsayat *et al*. showed that ensemble learning using different CNN architectures can improve ECG classification performance[5] . However, our proposed method streamlines the input data processing, as the ensemble members share similar architectures, unlike traditional ensembles, which typically combine models with different structures and input requirements.

Our study demonstrates that the format of ECG images used for training and classification is a critical factor in image-based deep learning approaches. When models were trained on either 4x3 or 2x6 images and tested on a different format (cross-format classification), there was a notable decline in performance compared to same-format classification. However, incorporating both formats into the training dataset, either by duplicating ECGs in both formats or by using a balanced mix, substantially mitigated this performance drop. As shown, classification performance using either the combined or mixed-format training datasets was comparable to that of same-format training and testing. We refer to the model trained on the multiformat dataset as *MultiECGNet*. MultiECGNet demonstrated robust performance across both ECG formats, achieving results similarto same-format classification while avoiding the performance degradation seen in cross-format testing. Overall, these findings suggest that training with a mixed– format ECG dataset, as done in MultiECGNet, effectively reduces the adverse effects of cross-format classification and supports more robust model generalisation.

Of note, full EfficientNet B3 model (model 385 in Figure 2) consistently demonstrated poor and low-confidence predictions during cross-format classification. This suggests that it may be more sensitive to layout-specific features tin ECG images. However, removing this model from the ensemble led to an overall decline in performance, as indicated by reduced F1 score. Therefore, despite its suboptimal performance in isolation, this model contributes complementary information that enhances the ensemble’s overall effectiveness.

Our models achieved comparable performance in atrial fibrillation classification to the results reported by Sangha *et al.*[2] , despite being trained on a much smaller dataset, 21,000 ECGs from the PTB-XL dataset compared to 2.7 million ECGs used in the Sangha *et al.* study. This suggests that our approach may be particularly valuable for studying less common conditions where large historical datasets are not available. An additional advantage of the ensemble learning approach is its flexibility: different models within the ensemble may exhibit varying sensitivity and specificity. This can be leveraged in clinical applications where specific performance characteristics are prioritised. For example, in scenarios requiring high sensitivity, such as ruling out myocardial infarction, a model favouring sensitivity over specificity may be more appropriate.

A key advantage of our technique is that an image-based approach can leverage extensive historical ECG databases, which have traditionally been archived as image files in medical records. While more recent ECG data are increasingly stored prospectively as digital signals, collecting a similarly large volume of high-quality digital ECG data for training deep learning models would take considerable time and resources. Furthermore, image-based ECG analysis offers practical benefits in certain clinical settings. For example, remote and regional health services may rely on smartphone camera-based ECG transmission[16] . As such, a robust image-based analysis pipeline remains highly relevant, even as digital signal acquisition becomes more widespread in prospective data collection.

An additional advantage of our approach is its potential applicability to other areas of medicine that rely on diagnostic imaging, such as pathology and radiology. For example, in a previous study, a single EfficientNet B3 model was trained on 149,130 histological images of various deep myxoid soft tissue lesions, achieving an accuracy of over 97% compared to 69.7% accuracy by pathologists[17] . Our ensemble-based approach could further improve performance in such contexts, particularly for conditions where only limited datasets are available.

### Limitations

There are several limitations to our study. First, the ensemble model approach is computationally more demanding than a single model, which may pose challenges when scaling to larger datasets. This requires increased computational resources and longer training times. However, future advancements in hardware and computing infrastructure may help mitigate this limitation.

Second, when evaluating the impact of ECG format on classification performance, we used only two formats, and both were equally represented in the training dataset. In real-world clinical settings, more than two formats may exist, often with an imbalance in their distribution, which may lead to a change in performance. Furthermore, we did not account for minor image rotations that are frequently present in clinical ECG images, such as those addressed in the Sangha et al. study[2] . While the presence of such rotations in both training and validation datasets may help mitigate this issue, it still represents a limitation in terms of real-world generalisability.

Finally, a key challenge that remains is the domain shift. That is, the reduced performance of models when applied to external or unseen datasets. This issue is common in machine learning and remains a major hurdle for the deployment of deep learning models in clinical practice. Developing methods to address domain shift would significantly enhance the generalisability and clinical utility of our ensemble approach.

## Conclusion

Our novel ensembling approach, which combines finetuned truncated variants of the EfficientNet B3 model, demonstrates improved performance over a single EfficientNet B3 model and achieves accuracy comparable to signal-based deep learning methods. Importantly, incorporating multiple ECG image formats during training enhances classification performance, underscoring the value of format diversity in image-based ECG analysis. This strategy supports broader applicability in clinical environments where ECGs are often stored or transmitted as images.

## Sources of Funding

This work was supported by an Australian National Health and Medical Research Council postgraduate scholarship (APP2022368) to JP, an Australian Medical Research Future Fund grant (APP2024443) to APH and a UNSW Cardiovascular and Metabolic Medicine seed grant to JP/JIV.

## Data Availability

All data produced in the present study are available upon reasonable request to the authors

## Notes

### Competing Interest Statement

The authors have declared no competing interest.

### Author Declarations

The study used data from the publicly available PTB-XL dataset, accessible via PhysioNet (physionet.org/content/ptb-xl/1.0.3/) and the China Physiological Signal Challenge dataset available at http://2018.icbeb.org/Challenge.html

